# Perspectives and Recommendations from Hospitalized Patients with Substance Use Disorders: A Qualitative Study

**DOI:** 10.1101/2023.09.16.23295657

**Authors:** Evan A. Balmuth, Sonali Iyer, David A. Scales, Jonathan Avery

## Abstract

**BACKGROUND:** Individuals with substance use disorders (SUDs) are hospitalized in growing numbers. Stigma is pervasive among their hospital providers, and SUD management during medical admissions is often inadequate. However, little is known about how these patients perceive their care quality. In particular, few studies have explored their positive care perceptions or recommendations for improvement.

**OBJECTIVE:** To explore perspectives on positive aspects, negative aspects, and consequences of care, as well as recommendations for improvement among hospitalized patients with SUDs.

**DESIGN AND PARTICIPANTS:** We conducted semi-structured, in-depth bedside interviews (*n* = 15) with patients who have been diagnosed with a SUD and were admitted to medical or surgical floors of an urban academic medical center.

**APPROACH:** Interviews explored patients’ hospital experiences and recommendations for improvement. The interviews were audio-recorded, transcribed verbatim, and imported into NVivo software. Two reviewers independently coded the transcripts using interpretative phenomenological analysis and inductive thematic analysis according to grounded theory, and recurring themes were identified from the data. Patients’ demographic and clinical data were analyzed with descriptive statistics.

**KEY RESULTS:** Perceived clinical and emotional proficiency were the most important components of positive experiences, whereas perceived bias and stigmatized attitudes, clinical improficiency, and inhumane treatment were characteristic of negative experiences. Such care components were most consequential for patients’ emotional wellbeing, trust, and care quality. Recommendations for improving care included specific suggestions for initiating and promoting continued recovery, educating, and partnering in compassionate care.

**CONCLUSIONS:** Hospitalized patients with SUDs often experience lower quality and less compassionate care linked to pervasive stigma and poor outcomes. Our study highlights under-recognized perspectives from this patient population, including socioemotional consequences of care and recommendations grounded in lived experiences. By striving to advance our care in accordance with patients’ viewpoints, we can turn hospitalizations into opportunities for engagement and promoting recovery.

## INTRODUCTION

The United States faces a worsening epidemic of substance use-associated morbidity and mortality. Over 46 million people had a substance use disorder (SUD) in 2021,^1^ and over 109,000 people died from overdoses in that year.^2^ Furthermore, an estimated 1 in 7 hospital admissions are for individuals with SUDs – and this proportion appears to be increasing over time.^3^ Despite the substantial number of patients with SUDs in hospitals, little is known about how they perceive the quality of their care.

Research on clinician attitudes and prescribing suggests that patients with SUDs face tremendous barriers to quality care in hospitals. First, stigma and negative attitudes toward substance use are pervasive among their providers, and tend to worsen through medical training.^4–8^ Physicians tend to reject the brain disease model of addiction, believing addiction is a choice that indicates moral failure.^6^ Second, largely due to a lack of knowledge and teaching, many hospital providers are uncomfortable treating SUDs – including opioid use disorder (OUD) and alcohol use disorder (AUD), for which evidence-based treatments are widely available.^4,9,10^ Together, these factors likely contribute to an established undertreatment of these SUDs in hospital settings, in addition to lower provider engagement, patient-provider collaboration, and treatment adherence.^10–17^

Considering this predominant stigma and discomfort in managing SUDs, it is not surprising that individuals with SUDs have reported negative experiences and mistrust in inpatient settings.^5,18–20^ Patients with SUDs have indicated that experiences of discrimination and stigma surrounding their SUDs, as well as undertreated withdrawal and ongoing cravings to use, are core drivers of their decisions to pursue patient-directed discharge (PDD) from internal medicine wards.^20^ Additional studies have revealed mutual mistrust between patients with SUDs and their hospital internal medicine providers, also influenced by stigma and negative preconceptions.^5,19,21^ This poses a substantial threat to care quality, as mistrust has been shown to jeopardize engagement in care and treatment adherence.^22^

In light of the challenges faced by the growing number of hospitalized patients with SUDs, a greater understanding of their perspectives and recommendations is needed. Only a small number of studies have explored these patients’ perspectives regarding their hospital experiences – and among these, even fewer have gathered their opinions on how care can be improved. Efforts to address these patients’ concerns and consider novel interventions can increase satisfaction, engagement, trust in the therapeutic alliance, and overall care quality.^5,10,12–17,19,20,22^ Furthermore, engaging patients in the evaluation and design of health services can enhance care delivery and education development.^23^

In this qualitative study, we explore perspectives and recommendations among hospitalized patients with SUDs through semi-structured interviews. Our approach is unique in assessing both positive and negative aspects, as well as consequences of care. In addition, we prioritize patients’ voices by highlighting their recommendations for improvement. Ultimately, greater appreciation of these often marginalized viewpoints can lead to more humane, higher-quality care for patients with SUDs – including novel interventions born from their lived experiences.

## METHODS

### Study Design

This is a qualitative descriptive study. We conducted semi-structured, in-depth interviews with patients during their hospital admissions at NewYork-Presbyterian/Weill Cornell Medical Center (NYP/WC), an academic hospital in New York City. The study was approved by the NYP/WC Institutional Review Board.

### Participants

Participants were drawn by convenience sampling from primary medical or surgical teams consulting the hospital’s addiction consult service (ACS). Eligible patients were 18 years or older and had a SUD diagnosis confirmed by chart review. Ineligible patients were those unwilling or unable to independently provide informed consent. A medical student interviewer (EAB) approached eligible patients at bedside, clarified that he was not part of their care team, explained the study’s purpose and confidentiality procedures, and requested verbal consent for participation in an approximately 30-to 60-minute audio-recorded interview. All but three approached patients were interviewed; those three expressed interest in participating, but deferred their decision and were discharged before interviews could be conducted. All participants received a $25 Amazon gift card.

### Data Collection

We obtained demographic and clinical characteristics (**Table 1**) by chart review. We conducted, audio-recorded, and transcribed 15 bedside interviews. Our interview guide was informed by prior studies involving patients with SUDs^19–21^ and incorporated novel questions exploring both positive and negative aspects, as well as consequences of care on patients’ experiences and their recommendations for improvement (**Supplemental Table 1**). We asked patients to reflect upon care in their present admission and prior admissions in any hospital. All patient and interview data were stored in REDCap.^24,25^

**Table 1.**
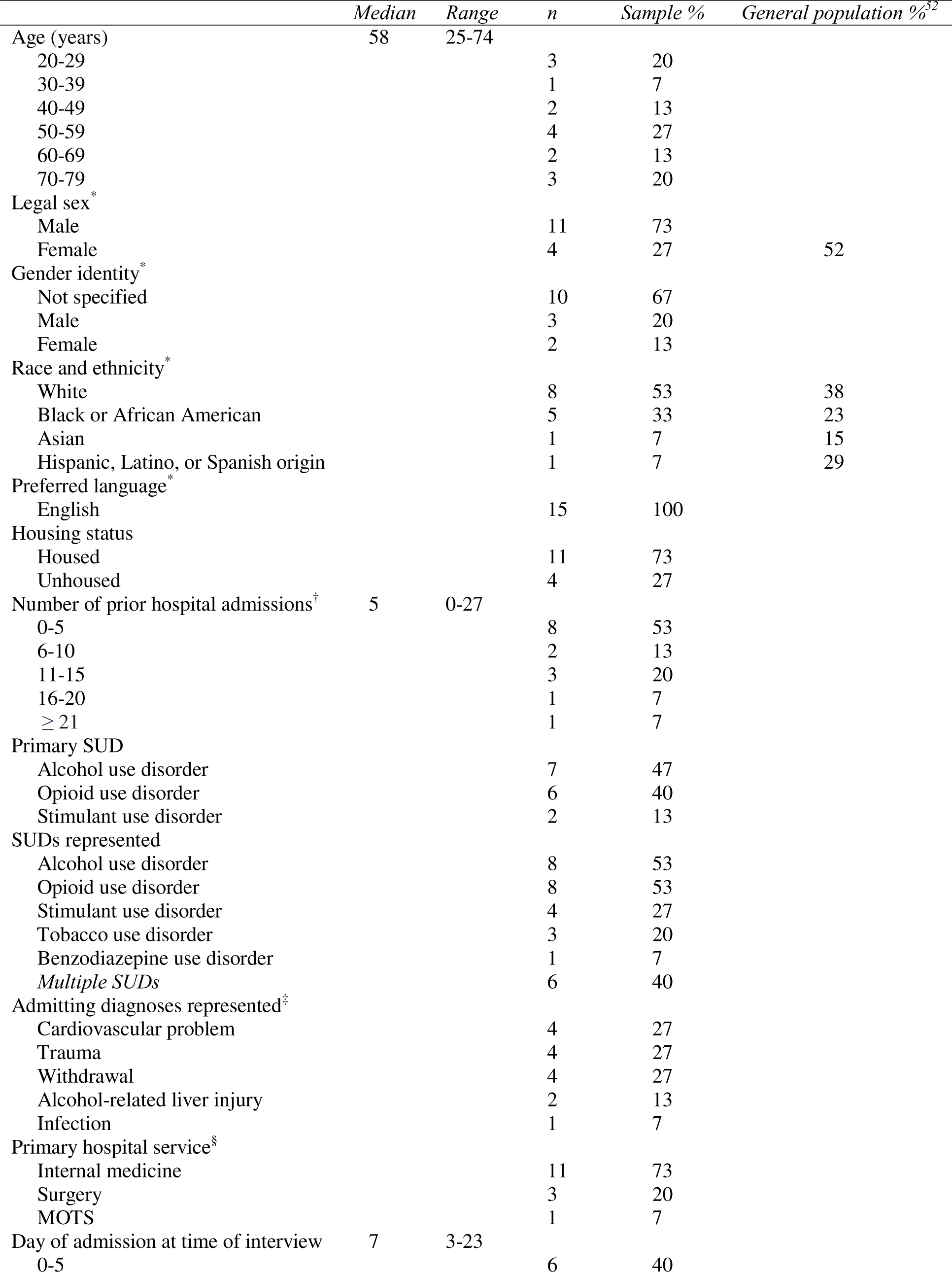
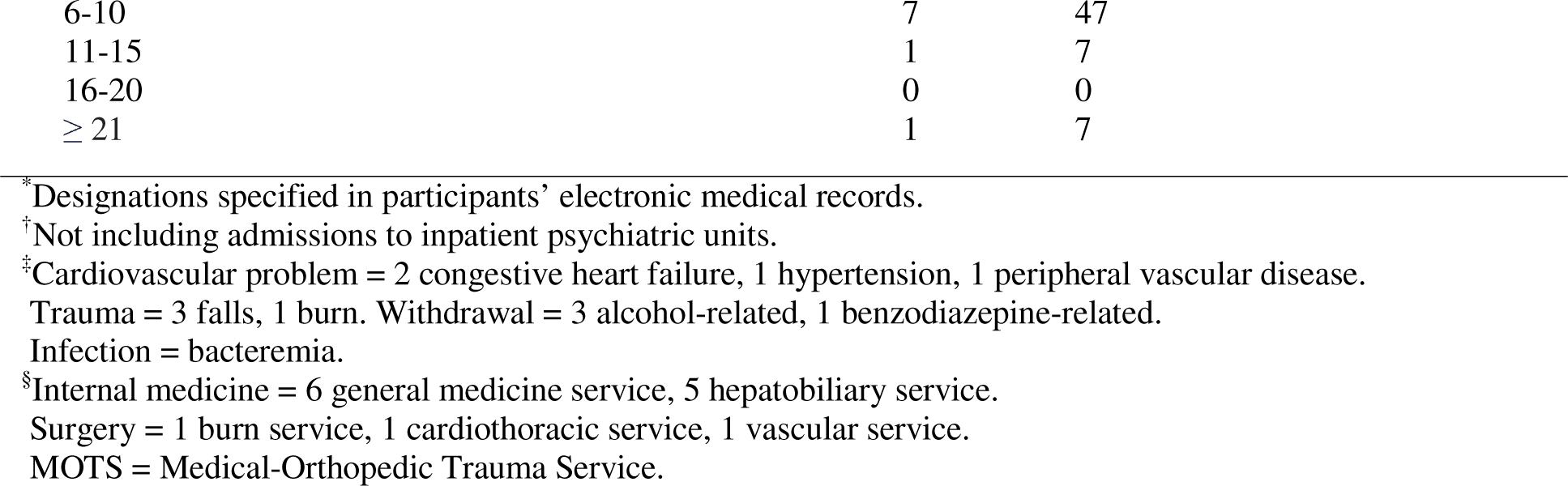
Demographic and clinical characteristics of study participants (*n* = 15).

### Data Analysis

Descriptive statistics were calculated in Microsoft Excel 16.73.^26^ EAB transcribed the interviews verbatim and uploaded the transcripts into NVivo 12 Plus.^27^ Two medical student analysts (EAB and SI) systematically coded the transcripts in NVivo^27^ through interpretative phenomenological analysis^28^ and thematic analysis,^29^ employing an inductive approach based on grounded theory to derive all themes from the data.^30^ Briefly, the analysts assigned codes to all new ideas identified in the transcripts; each code was applied to all recurrences of the same idea throughout its original transcript and all subsequent transcripts. The analysts cross-coded the first five transcripts, independently coding each transcript and regularly comparing codes toward consensus on a preliminary codebook. All subsequent interviews were coded separately. The analysts met regularly to discuss new codes, identify any occurrences of new codes in previous transcripts, and iteratively reorganize the codebook through a consensus-based discursive approach. After every five new transcripts were analyzed, the analysts reviewed their shared codebook to identify major and minor themes. Interviews were conducted in parallel to analysis, which continued until no new themes emerged from additional transcripts (theoretical saturation).^30–33^

Our COREQ qualitative research checklist^34^ is provided in **Supplemental Table 2**.

## RESULTS

### Demographic and Clinical Characteristics

**Table 1** shows the demographic and clinical characteristics of the study participants (*n* = 15) and New York City population. The median age was 58 years. Sex was 27% female. The majority of participants identified as “White” (53%) or “Black or African American” (33%) and carried a diagnosis of AUD (53%) and/or OUD (53%); 40% carried more than one SUD diagnosis.

### Perceptions of Care

**Table 2** lists the recurring themes of patients’ perceived positive aspects, negative aspects, and consequences of care. Where quoted, participants are identified by pseudonym initials.

**Table 2.**
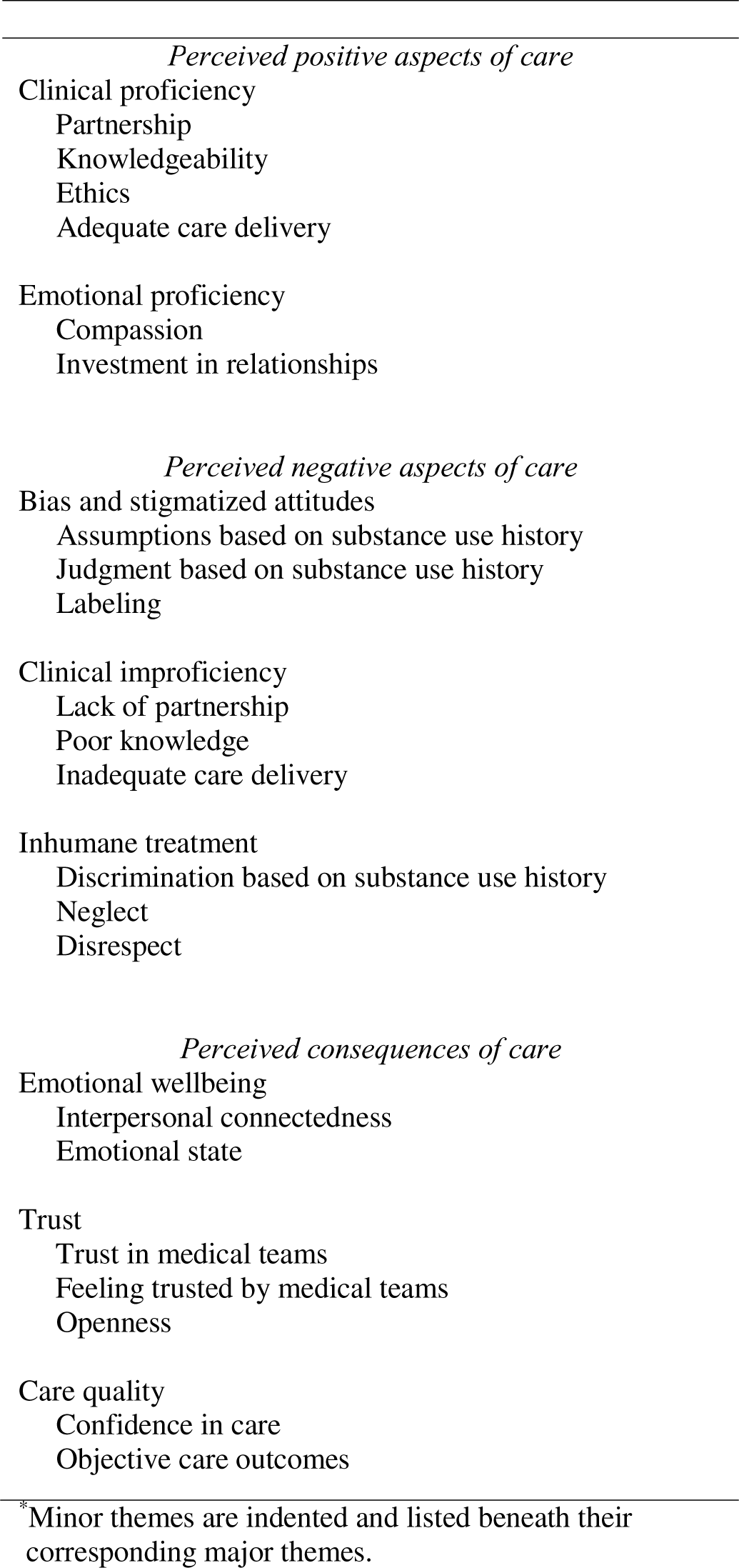
Recurring themes: patients’ perceived positive aspects, negative aspects, and consequences of hospital care.*

### Perceived Positive Aspects of Care

#### Clinical Proficiency

Within the domain of clinical skills, a provider’s ability to partner with their patient was seen as especially important. For example, most patients appreciated when providers listened to them, valued their input, and responded to their concerns. A.Z. described how happy and trusting she felt when the team promptly changed her withdrawal treatment after she indicated her symptoms were more due to opioid than benzodiazepine withdrawal:

> I was shocked … I was very happy … I’m like, “I’m happy that you listened, and you took my opinion into account.” Because, I clearly have knowledge in this area … Listening to what I’m telling you is going on in my body is important to me. I know that I trust that the people who are taking care of me trust me.

Individualized care was seen as another crucial aspect of partnership. B.Y. explained how she felt treated as an individual person, rather than stereotyped as an “addict” with all the associated stigma: “They actually treat you as the patient first, and being the addict as a secondary factor.” Similarly, C.X. expressed how motivated he felt when his team communicated in the way that worked best for him: “Everything that was told here really is … exactly what *I* wanted to hear. And how they make you feel when they see you pushing yourself, makes you want to push yourself more and more.” Several patients also appreciated when providers took a thorough substance use history to inform their care – such as D.W., who felt accommodated when the team asked about his non-prescribed methadone dose and tailored his pain regimen accordingly: “I let them know that I was using in the street, and they asked me what was the dosage I was using. And I told them, and they accommodated me.”

Many patients also praised providers’ knowledgeability – especially regarding SUD management. For example, E.V. commended his addiction medicine providers for their expertise as trained specialists: “They know their stuff – they’re specialists in the field, you know. Especially the addiction specialists.” From another standpoint, B.Y. felt most trusting of a provider whose knowledge was informed by personal experiences with addiction: “They had been there … Sometimes – unless you’ve experienced it either firsthand or through watching a family member go through it – there are certain things you just don’t understand.”

In addition, patients emphasized how providers upheld ethical codes of conduct. Most praised their teams for providing equal care regardless of substance use history, and for giving their best effort. For example, F.T. stated: “They don’t treat you any different here. If you’re an addict here, they do everything they can to make you comfortable.” Many patients, including F.T., also believed their providers had good intentions and cared deeply: “I truly believe that they have the best interests of their patients at heart.” Some, including E.V., appreciated when their providers not only intended to treat their most immediate concern, but also lead them on a path to long-term recovery: “I think they’re very sympathetic and want to help you – they want you to quit.” “They’re hoping that their advice leaves a lasting impression on you.”

Perceptions of adequate pain control and SUD treatment were also key to a positive experience. For example, F.T. appreciated receiving analgesics as needed, without suspicion of ulterior motives: “If … I called and needed pain medication, I got it, no questions asked. There was none of that, ‘are you drug seeking right now?’” G.S. explained how, after expressing his goal of abstinence, a prior hospital team helped him achieve it by starting medications for OUD: “They worked with me, and gave me all the proper medicines and … being motivated physically, spiritually, mentally – I kicked all that heavy drug stuff.” For many, efficiency and speediness were seen as fundamental elements of effective care delivery. As G.S. expressed: “I really love the way everyone works here so efficiently … When you ask something – not that it be right away, but … as soon as possible … They always come through here.”

#### Emotional Proficiency

Alongside components of clinical aptitude, perceived emotional proficiency was also central to positive experiences. Specifically, patients emphasized the importance of treating with kindness and compassion. For example, B.Y. remarked: “It’s been wonderful. The teams here have always been so nice to me;” H.R. stated: “Kindness is the biggest thing.”

Beyond taking time to discuss clinical matters, many appreciated when their providers invested in building a relationship. I.Q. experienced that trust improved when providers got to know him on a personal level: “The better they get to know me, the better the trust.” J.P. explained that humor and small talk can lead to more openness, trust, and even friendship through a prolonged admission:

> [The provider] will make a joke with me. Maybe she said something about her family for example, or vacation … and then open up a little bit. I say, “oh, yeah, I was there…” And you build the trust, and then you can, [over the course of] four months, [build] the friendship.

Indeed, some patients developed particularly strong connections with their providers. For example, F.T. was especially moved by the encouragement he received from an addiction medicine physician:

> Dr. X was the first person in my lifetime … to sit across from me and look me in my eyes and tell me that she believed in me … and that she – as of knowing me for a couple weeks now and talking to me every day – she fully believed in me 100 percent that I could … get past the withdrawal and … not only can I get sober, but that I can maintain my sobriety for my lifetime … That meant a lot to me.

### Perceived Negative Aspects of Care

#### Bias and Stigmatized Attitudes

Most patients had experienced stigma in hospitals, and perceived that this often took the form of assumptions based on their substance use history. B.Y. described how people with SUDs are often assumed to be “drug-seeking,” leading to lower quality care, neglect – and sometimes, unnecessary worsening of illness and death: “A lot of times, a lot of people get really sick and die because they were treated with subpar care or weren’t treated with care at all, because they think that we’re drug-seeking.” She elaborated that many providers seem reluctant to provide pain medication based on the assumption that this will worsen the SUD. However, without adequate pain control, she might be inclined to pursue PDD:

> I think it’s like, they think that because I’ve done drugs, if they give me something for pain that it’s gonna trigger me to do something worse. When in reality … if I’m not being medicated to a point that I’m comfortable, it makes me want to leave.

K.O. explained how she felt labeled by her AUD diagnosis, which follows her across hospitalizations and leads to perceived judgment:

> I think you get labeled … You know as a kid they would tell you, “that’s going on your permanent record!” That’s how you feel. Like, this is gonna follow you. Everywhere you go … I think people view you differently. I think that there is judgment.

#### Clinical Improficiency

A lack of partnership was often seen as detrimental to care quality. Several patients described a sense that they were treated with a uniform approach for all people with their SUD – even if it was not the right fit. For example, K.O. remarked: “Everybody’s very different. We’re all individuals, so our problems are gonna be individual … It just seems like there’s just this one blanket answer for everything.” She elaborated that teams too often pushed for abstinence, without asking patients what *they* were ready for and what the implications of that change would be. As a result, she felt blamed for what she considered to be an important part of her identity:

> I think it’s a personal choice, and to be made to feel almost like you’re a failure… But you’re not a failure … it’s a part of your life … it’s part of who you are. And if it’s, “you’re sick! You’re sick!” But it *is* who you are.

Such “one-size-fits-all” approaches also impeded trust. As A.Z. explained: “If a doctor comes in and this is … the, ‘you’re ok, trust me, I’ve seen a hundred cases like this’ doctor; you can’t trust a doctor like that, because every human is an individual.”

Partnership was also hampered when patients felt their personal experiences with substance use were not respected. For example, K.O. felt distrusted when the team did not seem to believe her own explanation for her anxiety symptoms, which she found most consistent with a panic attack; instead, the team insisted it was alcohol withdrawal – despite the patient’s extensive experience with both conditions. Furthermore, she felt frustrated with her medical team’s apparent knowledge gap – she knew it was unlikely for withdrawal to play a major role after one week in the hospital:

> I was like, “ … I’m having an anxiety attack, this is not withdrawal” … Can’t you just believe me? … I’m not lying … I’ve been drinking a very long time. I’ve done the withdrawal on my own … We all know after a week I’m not really gonna have … a seizure or a whatever.

Perceived inadequate care delivery was another primary concern. Many felt they did not have enough time or familiarity with their providers. F.T. explained how this impeded trust-building: “I don’t know them well enough. They haven’t spent enough time with me for me to turn around and say I trust them.” Waiting to receive care was also viewed as a common problem. E.V. expressed how frustrating it can be to wait for medication when experiencing withdrawal: “When … you’re going through withdrawals, and you expect to get immediate medication … sometimes it takes a while for them to order it, or get it and so forth, and it sort of irritates you.” Inadequate pain and withdrawal management were also common concerns. G.S. described how he was not discharged with enough pain medication after surgery – and this almost triggered a return to use:

> When I got home I realized … I’m in so much pain … with all the staples, and with only like a week’s supply – not even – of the oxys … I’m in pain now, and I’m already down to a few of these things … That was the scariest moment of my life … because the fact of just going home, what’s gonna happen – especially people like myself – you know, ex-addicts, it’s easy for us to just go pick up a bag of heroin.

L.N. described a time when he was hospitalized after an opioid overdose, but was not treated to prevent withdrawal: “When I overdosed in the past, I’d be in the hospital and then … you’d be there for a day and then you can get withdrawals while you’re there.”

#### Inhumane Treatment

Treatment was frequently perceived to cross a line from low quality to inhumane. For instance, patients described numerous experiences of discrimination based on their substance use history. This often centered on issues of pain control; F.T. explained that revealing a SUD diagnosis often results in inadequate pain management:

> A lot of times, when you’re an addict and you come into the hospital, they’re not liberal with pain medication when you tell them you’re an addict. It doesn’t benefit to be truthful with them about those things in hospitals, unfortunately, when it should.

Discrimination was also perceived in the form of dismissing medical concerns. B.Y. described an experience with a physician who she felt was judgmental, did not take her complaints seriously, and would have missed a diagnosis had she not requested a different provider: “I actually had a severe underlying condition that would’ve gone completely unnoticed if I had stuck with the original physician put on me because he basically writes off … anybody who has addiction issues.” B.Y. additionally perceived that patients with AUD may be treated better than those with OUD:

> I feel like they take alcohol use way more serious. They care way more about it. Like if you have a history of alcohol use, they’re gonna make sure that you’re more comfortable than an opioid user because there’s more clinical evidence that alcohol withdrawal can kill you.

Several patients also reported experiencing complete neglect. F.T. described an instance when he sought withdrawal treatment, but never received care:

> I told them the situation, the actual situation, that I have money, I just can’t find anything, and I’m really getting sick and desperate, and I need help. And they let me sit in that waiting room – I’d still be sitting there now if I didn’t just get up and leave. They never called me again … They just pretended and acted like I just straight up did not exist.

In addition, patients reported frequent experiences of blatant disrespect from providers and other hospital staff. This took many forms, from rude comments to taunting and even physical aggression.

A.Z. described the experience of another individual with a SUD who she accompanied to the hospital:

> They treated him disrespectfully, they … handcuffed him when they didn’t know what to do with him. They were physically violent with him, they spoke to him disrespectfully, and, first and foremost, they didn’t help him. And they kind of made fun of him and taunted him, like, “oh, he’s doing that thing again.” And it was very upsetting.

B.Y. explained that she sometimes feels blamed for her acute medical conditions because of her SUD history:

> At another hospital, they would have just told me to shut up and that I’m just a big baby and that I did it to myself. And that if I didn’t want to feel this way then I shouldn’t have done drugs. Nobody deserves to be treated like crap just because they got high.

### Patient Recommendations

**Table 3** lists the recurring themes of patients’ recommendations for improving care, alongside examples of specific initiatives they proposed and supporting quotations. One recurring theme among patients’ recommendations was to initiate and promote continued recovery; specific suggestions included psychotherapy, publicizing and offering to initiate SUD treatment, helping patients prepare for exposure to substance-related cues post-discharge, and establishing post-discharge pain management plans. Another theme centered on education – specifically, augmenting SUD education for both patients and providers. A third theme was promoting partnership in compassionate care; suggested approaches included treating with compassion, individualizing care, prioritizing patient viewpoints and increasing accountability through satisfaction surveys, and listening to both patients and their advocates.

**Table 3.** Recurring themes: patients’ recommendations for improving hospital care.

| *Recommendation* | *Example initiatives* | *Example supporting quotations\** |
| --- | --- | --- |
| Initiate and promote continued recovery | Offer group therapy (inpatient and outpatient) | *A couple of times they had group talks ... The people that went got something from it. Maybe if you promoted that a little more, and just let people chat it out ... [Before going] to the AA meetings, I was like, "man, my life sucks." And then you go to AA meetings and you're like, "oh my God, people are a lot worse" ... When the meeting was over ... you start a conversation with an addict, and then, if you become friends with an addict and they both want to stay together, then you figure out ways to do it. (M.U.)*<br>*I think there should be papers in [patient rooms] saying, "are you having trouble with a struggle with substances that you're scared to bring up? This is a number that's a safe place to call. You will not get in trouble for calling. If you just want to know what detoxing would look like, just call this center." (A.Z.)* |
|  | Offer individual psychotherapy |  |
|  | Post flyers publicizing compassionate SUD treatment |  |
|  | Practice "real-world" re-immersion |  |
|  | Offer to connect with post-discharge SUD treatment |  |
|  | Make a post-discharge pain management plan |  |
| Educate | Teach patients about SUD and its consequences | *So much of suicidal attempts start out as just being so hooked on drugs that the road to recovery looks too difficult ... It needs to be publicized that there are ways to make withdrawal easier, that detoxing is not going to be as hard as you think, and there are a lot of people who have walked this path before you – and they'll light the way for you. (A.Z.)*<br>*I think that [medical trainees] really need to know more about it ... Is there a chapter in a book somewhere about this? You know, I think they should have to deal with people for a while – with a problem ... I think it should be part of a rotation for them ... Somewhere in their training there, they need to know more. (K.O.)* |
|  | Augment addiction medicine training for providers |  |
| Partner in compassionate care | Treat with compassion | *By just being kind, you know. Addiction is a disease ... A lot of people ... they're doing drugs to escape a feeling of being abused ... Yes, kindness is the biggest thing. (H.R.)*<br>*They have a questionnaire that you fill out, asking ... how was you treated from 1 to 5 ... It matters because that survey is taken by everybody ... it makes a difference for the staff. Because they know that they're gonna be viewed by it, so they want to do a good job ... So I know that I'm gonna have a better off experience, because they don't want to be reprimanded for the way I was experienced. (D.W.)* |
|  | Individualize care |  |
|  | Distribute patient satisfaction surveys |  |
|  | Listen to patients and their advocates |  |

\*Initials representing the quoted participant's pseudonym are included in parentheses after each quotation.

## DISCUSSION

This qualitative study presents crucial perspectives from hospitalized patients with SUDs, notably emphasizing the under-explored domains of perceived positive aspects and consequences of care as well as recommendations for improvement. Whether expressed through positive or negative experiences, patients overwhelmingly underscored elements of partnership – with providers valuing their input and choices – as core components of their expectations for clinical proficiency. Meeting these expectations is key to helping patients onto a path of recovery. Hospitalized patients with SUDs have previously highlighted the importance of choice and patient-centered, compassionate, non-judgmental care for building trust and readiness to change.^19–21,35^ Our findings elaborate upon prior work by describing how patients often feel treated with a uniform approach, and emphasizing the importance of individualized care informed by the patient’s readiness to change. Effective partnership in this setting undoubtedly requires knowledgeability and practice on the part of providers. Previous studies have demonstrated central roles for provider knowledgeability and expertise in managing SUDs, as perceived by their patients; these have been shown to influence patients’ trust and readiness to engage in treatment.^19,21^ Our study expands upon this work by highlighting that personal experience with addiction can put certain team members in a unique position to build trust. This is consistent with research suggesting that peer mentors – individuals currently in recovery who offer to share their lived experiences – hold great potential for improving hospital care.^35^

Our participants also perceived stigma to be pervasive in hospital settings, yielding profound repercussions. Specifically, patients reported feeling stereotyped based on the “label” of SUD in their medical history, in addition to stigmatized assumptions that they were “drug-seeking” or that adequately treating pain could precipitate relapse – prejudices which have been previously described.^20,36,37^ Our findings additionally support a body of work showing that hospitalized patients with SUDs experience bias and discrimination in their care – often manifesting with inadequate pain and withdrawal management.^5,19–21^ One participant perceived preferential treatment for patients with AUD compared to those with OUD, especially regarding withdrawal management; this may reflect prior work demonstrating more negative resident attitudes toward patients with OUD than those with AUD,^38^ though further study is needed to better understand how experiences vary among patients with different SUDs. Regardless of the specific SUD diagnosis, stigma, bias, and discrimination have far-reaching consequences for the patient-provider relationship and downstream outcomes. For our participants, trust in their providers, as well as the degree of openness and honesty in the therapeutic alliance, were especially dependent on their perceptions of the care they received. In addition, some aspects of care – such as feeling that medical concerns were dismissed – affected patients’ own sense of trustworthiness. Participants also provided unique insights into emotional consequences of their care; often illuminating the fear and despair that can result from poor treatment, they also highlighted the inspiration and positive outlooks that can arise from compassionate care. Our findings build upon prior work which has demonstrated lower treatment adherence and engagement in care among patients with SUDs who experience stigma and discrimination.^4,11,12,14^ Furthermore, pain and withdrawal management have previously been identified as important contributors to trust, readiness to change, and – when inadequately addressed – decisions to pursue PDD.^20,21,36^ Prior studies have also shown that hospital providers tend to lack knowledge and comfort in managing SUDs, and that withdrawal tends to be undertreated for hospitalized patients with OUD.^10,17^ Moreover, persistent pain has been associated with relapse and poor outcomes in outpatient SUD treatment,^39,40^ though a role for undertreated pain following medical hospitalizations remains under-appreciated. Further study is needed to understand providers’ biases and challenges regarding pain control for hospitalized patients with SUDs, and to fully characterize the scope of undertreated pain toward evidence-based care improvements.

Patients imparted salient recommendations for improving their hospital care which are substantiated in existing literature and warrant further investigation. For example, many recommendations aimed to facilitate partnership in compassionate care through individualization and listening. This approach parallels principles of motivational interviewing: an evidence-based, patient-centered method for enhancing intrinsic motivation and guiding care based on the patient’s values and readiness to change^41^ which requires further study in inpatient settings. Psychosocial interventions, including group and individual psychotherapy, were also recommended. Various types of psychotherapy have established evidence bases in outpatient and psychiatric inpatient settings.^42,43^ A small body of work suggests psychotherapy can reduce readmission rates after medical hospitalizations;^44^ however, future studies will need to determine which approaches are most effective in hospital settings. Participants also recommended key interventions – such as ensuring a post-discharge pain management plan and connecting patients with timely post-discharge SUD follow-up – that could improve safety during the days following hospital discharge, which represent a high-risk period for return to use and overdose.^45^ Finally, patients recommended augmenting SUD education. Given the established knowledge gaps among hospital providers, there remains a substantial need to expand SUD education at all levels of medical training.^9,10^ Some initiatives have demonstrated effectiveness in improving knowledge and confidence managing SUDs, as well as reducing stigma among residents;^38,46,47^ however, such programs are not widely implemented. Participants also recommended more in-hospital SUD education for patients, especially surrounding substance use-related health consequences and treatment options. During medical admissions, SUD management is often deferred to outpatient follow-up; however, studies have established the feasibility of initiating treatment for AUD and OUD prior to discharge.^48–50^ Our study builds upon this work by emphasizing that many patients with SUDs may be eager to learn about, initiate, or resume treatment during medical admissions; for those who are not yet ready for treatment, the admission is an opportune time to provide education as an initial step toward recovery.

Our findings should be considered in the context of certain limitations. First, participants could have tailored responses to avoid offending their providers; we aimed to mitigate this potential courtesy bias^51^ by asking participants about experiences across hospitalizations, emphasizing that the interviewer was not involved in their care, and underscoring that their responses would be anonymous and not shared with their care team. Reassuringly, nearly all participants discussed both positive and negative experiences; interviews tended to be neither overwhelmingly positive nor negative. Second, this study was conducted in a single urban academic medical center and utilized convenience sampling. Although responses covered experiences across numerous hospitals, their generalizability remains limited. It is imperative that future studies explore variations in patient perspectives across SUD sub-types, clinical settings, and underrepresented backgrounds.

Amidst pervasive stigma and a growing burden of substance use-associated morbidity and mortality, hospitalized patients with SUDs often experience lower-quality and less compassionate care linked to poor outcomes. Our study is one of the first to engage these patients in assessing their hospital care and producing recommendations for improvement. By amplifying their voices, we aim to raise awareness of care inequalities and downstream repercussions alongside innovative recommendations grounded in their lived experiences; these should inform future studies and novel interventions toward better care for individuals with SUDs. Our findings suggest that ensuring patients feel heard by meeting their expectations for clinical and emotional proficiency should yield great potential for improving health. Follow-up studies must further explore these expectations, approaches to meeting them, and resultant outcomes. Ultimately, by striving to advance our care in accordance with patients’ viewpoints, we can turn hospitalizations into opportunities for engagement and promoting recovery.

## Data Availability

All data produced in the present study are available upon reasonable request to the authors.

## Acknowledgements

The authors thank the Weill Cornell ACS for helping us identify potential participants and providing exceptional care to hospitalized patients with SUDs; the Weill Cornell Area of Concentration (AOC) Program for fostering research mentorship opportunities; the Cornell Center for Social Sciences for assistance with NVivo software; and all the patients who offered their time and insights for this study.

## Funding Information

This study was supported by Weill Cornell Fund 61500693.

## Compliance with Ethical Standards

This study was approved by the NYP/WC Institutional Review Board (22-08025196).

## Conflicts of Interest

The authors report no conflicts of interest.

## SUPPLEMENTARY APPENDIX

**Supplemental Table 1.**
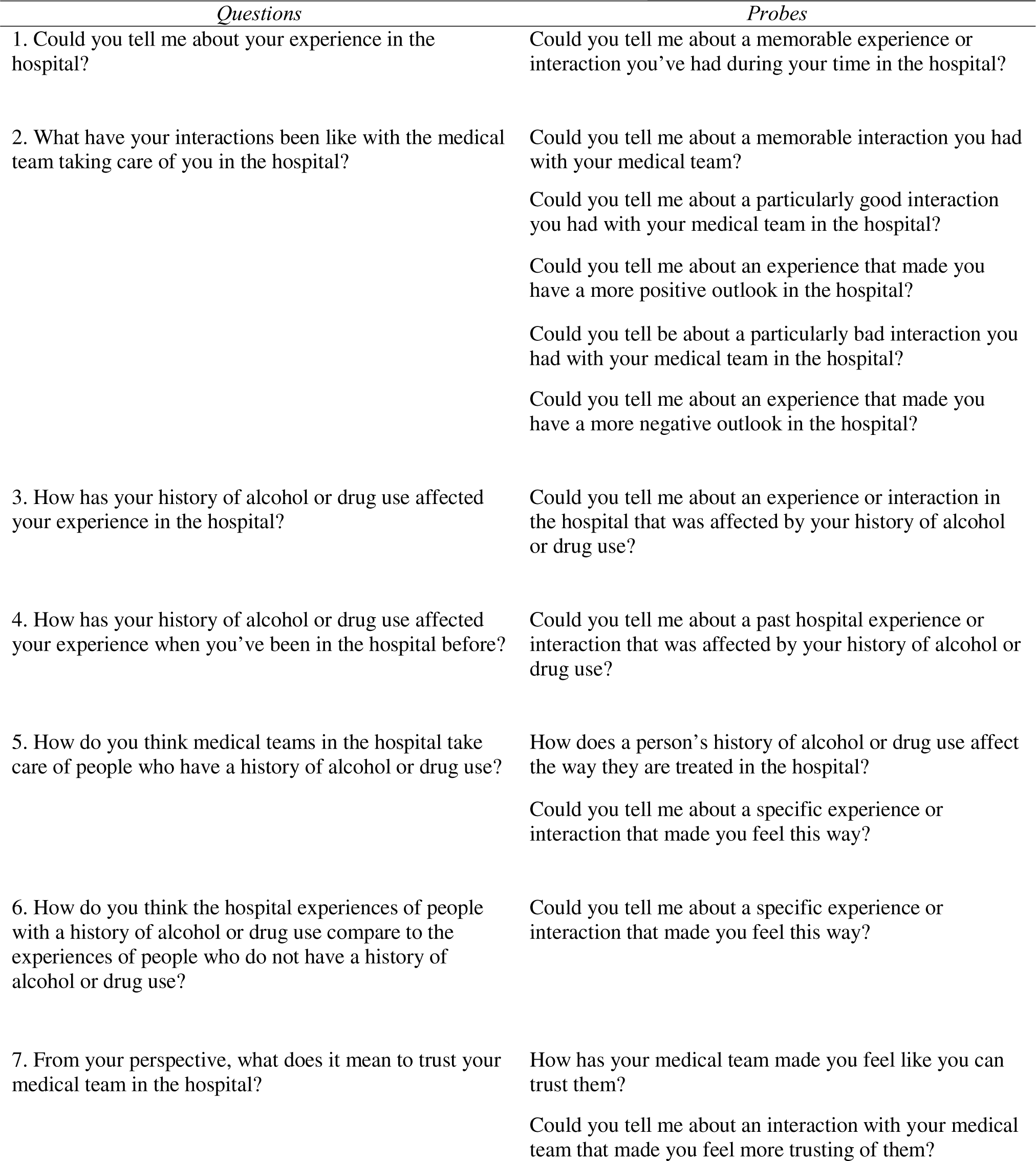
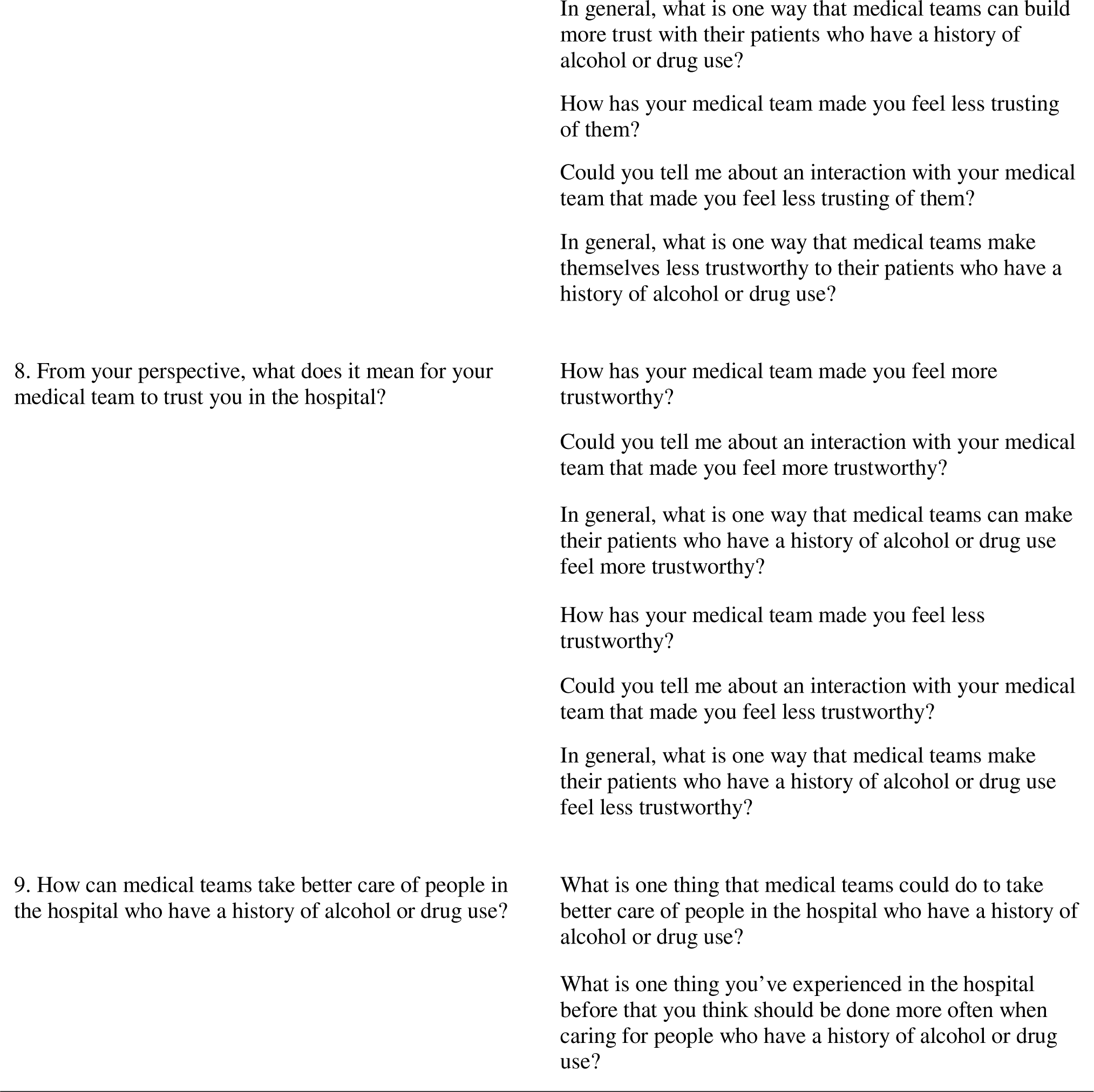
Semi-structured interview guide.

**Supplemental Table 2.** COREQ checklist for qualitative research^34^ applied to the present study.

| *Domain* | *Item* | *Description* |
| --- | --- | --- |
| Research team and reflexivity |  |  |
| Personal characteristics | 1. Interviewer/facilitator | EAB |
|  | 2. Credentials | B.S. |
|  | 3. Occupation | Medical student |
|  | 4. Gender | Male |
|  | 5. Experience and training | Qualitative methods, care of patients with SUDs, anti-stigma patient-centered communication |
| Relationship with participants | 6. Prior relationship | None |
|  | 7. Participant knowledge of the interviewer | Medical student at NYP/WC not involved in participants' care, researching experiences of patients with substance use history |
|  | 8. Interviewer characteristics | Project conceptualized for MD program research course, informed by interests in addiction medicine and internal medicine |
| Study design |  |  |
| Theoretical framework | 9. Method orientation/theory | Grounded theory and phenomenology |
|  | 10. Sampling | Convenience |
| Participant selection | 11. Method of approach | Face-to-face |
|  | 12. Sample size | 15 |
|  | 13. Non-participation | 3 deferred decision and were subsequently discharged |
| Setting | 14. Data collection setting | In hospital (NYP/WC), at bedside |
|  | 15. Presence of non-participants | Hospital roommate present behind separation curtain; occasional brief interruptions by treatment team for routine medical care |
|  | 16. Sample description | See Table 1 |
| Data collection | 17. Interview guide | Based on prior studies;<sup>19-21</sup> see Supplemental Table 1 |
|  | 18. Repeat interviews | None |
|  | 19. Audio/visual recording | Audio recording |
|  | 20. Field notes | None |
|  | 21. Interview duration | 20 minutes (median), range 6-53 |
|  | 22. Data saturation | Interviews and analysis conducted until theoretical saturation<sup>30-33</sup> |
|  | 23. Transcripts returned | No |
| Analysts and findings |  |  |
| Data analysis | 24. Number of data coders | 2 (EAB and SI) |
|  | 25. Coding tree description | See Methods section for description of coding approach |
|  | 26. Derivation of themes | Derived from the data |
|  | 27. Software | NVivo 12 Plus<sup>27</sup> |
|  | 28. Participant checking | No |
| Reporting | 29. Quotations presented and identified | See Results section and Table 3; participants identified by pseudonym initials |
|  | 30. Data/findings consistent | See Results and Discussion sections |
|  | 31. Clarity of major themes | See Results section, Table 2 and Table 3 |
|  | 32. Clarity of minor themes | See Results section, Table 2 and Table 3 |

## Notes

### Competing Interest Statement

The authors have declared no competing interest.

### Author Declarations

The IRB of Weill Cornell Medicine gave ethical approval for this work.

### Summary of Updates

Introduction, Methods, Results, and Discussion sections updated for conciseness; Supplemental Table 2 added.

